# Distribution of gestational age by maternal and infant characteristics in US birth certificate data: informing gestational age assumptions when clinical estimates are not available

**DOI:** 10.1101/2022.10.19.22281268

**Authors:** Andrea V Margulis, Brian Calingaert, Alison T Kawai, Elena Rivero-Ferrer, Mary S Anthony

**Author notes:** **Corresponding Author:** Andrea V Margulis, RTI Health Solutions, Av. Diagonal, 605, 9-1, 08028 Barcelona, Spain. **Previous Presentations:** Part of this work has been presented as a poster at the 2022 International Conference on Pharmacoepidemiology & Therapeutic Risk Management (24-28 August 2022; Copenhagen, Denmark). **ETHICS REVIEW STATEMENT** The RTI International institutional review board reviewed the protocol and determined that the study did not constitute research involving human subjects (RTI IRB STUDY00021950).

## Abstract

We aimed to describe the distribution of gestational age at birth (GAB) to inform the estimation of GAB when clinical or obstetric estimates are not available for perinatal epidemiologic research. We estimated GAB (median, mode, mean, standard deviation) and percentage born at each gestational week in groups based on plurality and other variables for live births in CDC’s US birth data.

In 2020, 3,617,213 newborns had birth certificates with nonmissing GAB. Among singletons (3,501,693), median and mode GAB were both 39 weeks. Births with lower median GAB were from women with eclampsia (37 weeks) or receiving intensive care (37 weeks); newborns receiving intensive care (37 weeks); infants with birth weight < 2,500 grams (35 weeks), < 1,500 grams (28 weeks), or < 1,000 grams (25 weeks); and newborns not discharged alive (23 weeks). Among twins (112,633), median GAB was 36 weeks (mode, 37 weeks). Additional noteworthy groups were women with 7-8 (median, 35 weeks) or 0-6 prenatal visits (median, 34 weeks) or aged 15-19 years (median, 35 weeks).

Some maternal and infant groups had distinct GAB distributions in the US. This information can be useful in estimating GAB when individual-level clinical estimates are not available.

## INTRODUCTION

In observational studies, researchers who use existing data sources to ascertain medication use or other exposures or events in pregnancy need to know when each pregnancy in the study population started. Whenever possible, researchers use obstetric or clinical estimates; otherwise, they typically use coded information available in their data. *International Classification of Diseases, Tenth Revision, Clinical Modification* (ICD-10-CM) Z3A codes for gestational age facilitate the process in claims data sources in the United States (US) to a certain extent.^1^ For pregnancies without the relevant clinical or obstetric information and without informative codes, researchers use other estimation methods. Often, such pregnancies are assigned a fixed duration based on the observed mode or median gestational age at birth (GAB) of pregnancies with some known characteristic, such as 34,^2^ 35^,1,3,4^ or 36^5^ weeks for preterm live births; 39^1,4^ or 40^2,3,5^ weeks for term live births, or 37 weeks for multifetal pregnancies.^5^ Then, the assigned GAB is subtracted from the delivery or birth date (which is usually available) to estimate the pregnancy start date and assess the timing of exposure relative to pregnancy start.

Healthcare claims and electronic health records contain information that might be used to identify groups of pregnancies with specific characteristics for which the GAB distribution differs from that of the general population of pregnancies; these distributions can in turn be used to estimate pregnancy start more accurately in those groups. The objective of this work was to describe the distribution of GAB in US birth certificates in groups defined by maternal or newborn characteristics that may also be captured in US healthcare claims or other data sources—e.g., plurality (singleton, twin, etc.), maternal age, race/ethnicity, smoking during pregnancy, body mass index (BMI) categories, birth weight—to inform the estimation of GAB when clinical or obstetric estimates are not available.

## METHODS

The completed checklist for methods reporting in perinatal pharmacoepidemiology^6^ is presented in Appendix A, Table A-1.

### Data source

We used US birth data files of the Centers for Disease Control and Prevention (CDC)^7,8^ for years 2019 (the most recent year before the COVID-19 pandemic) and 2020 (the most recent available data). These files are publicly available for download. Each row corresponds to 1 live birth and contains information on the mother, the pregnancy, and the offspring. We included live births to foreign residents^9,10^; this is why our totals are about 0.25% larger than the ones in CDC’s final reports, which did not include these live births.^11,12^ Variables correspond to the fields in US birth certificates; GAB is an obstetric estimate. Fields that might facilitate identification of individuals are not included in the downloadable data. No linkage with other data sources was sought in this study.

### Study population

The study population included all pregnancies ending in a live birth with nonmissing GAB; pregnancies with fetuses with chromosomal abnormalities or congenital malformations (minor or major) and fetuses from multifetal pregnancies were included. Women may have contributed pregnancies in 1 or both years; this information is not directly available from the data source; intrafamily correlation was not considered in the analyses. Each analysis included pregnancies with nonmissing values for the variables used in that analysis.

### Variables

Study variables are listed in Appendix A, Table A-2. Variables for this study were a subset of the variables included in the data source.^9,10^ GAB is provided as the number of completed weeks at the time of birth (range, 17-47).

### Statistical analysis

The unit of analysis in this study was live births, but, for clarity, maternal or pregnancy characteristics were described in terms of women or pregnancies; offspring characteristics were described in terms of newborns.

We estimated summary statistics for GAB and percentage of infants born with each gestational week (e.g., 0.1% born with 17 weeks, 0.2% born with 18 weeks) in various groups of live births, separately for 2019 and 2020. Each analysis included pregnancies with nonmissing values for the variables used in that analysis. GAB distribution is presented for all live births, for singletons only, and for twins only. Because these tables are sizable, the complete tables are presented with the supplemental information (see Appendix B: Table B-1, results for all live births; Table B-2, results for singleton live births; Table B-3, results for twin live births).

To explore whether the median, mode or mean would result in a smaller estimation error, we calculated 2 metrics for each of the 3 summary statistics: the mean squared error and the mean absolute value of the error. The mean squared error using the median in group X (e.g., singletons born small for gestational age) was calculated as follows: the observed GAB for live birth *i* in group X minus the median GAB in group X, squared, averaged across all newborns in group X. Similar calculations were conducted for the mean and mode. The mean absolute value of the error was calculated similarly, applying the absolute value instead of the square. Smaller mean squared error or mean absolute value of the error reflect a more precise estimation. More details on the methods are presented in Appendix A and results are presented in Appendix B, Table B-4.

## RESULTS

### Overall

In 2019, 3,757,582 live born infants were issued birth certificates in the US; 3,755,044 (99.9%) birth certificates had information on GAB (Table 1 and Appendix B, Table B-1). Median GAB was 39 weeks; mode, the same; and mean (standard deviation [SD]), 38.4 (2.1) (Appendix B, Table B-1). In 2020, there were 3,619,826 live births; 3,617,213 (99.9%) had information on GAB (Table 1 and Appendix B, Table B-1). The GAB median, mode, mean, and SD were nearly the same as in 2019 (Appendix B, Table B-1). Results for 2019 and 2020 were very similar (Table 1 and Figure 1); for further descriptions, we use data from 2020, the latest available information at the time of study conduct.

**Table 1.**
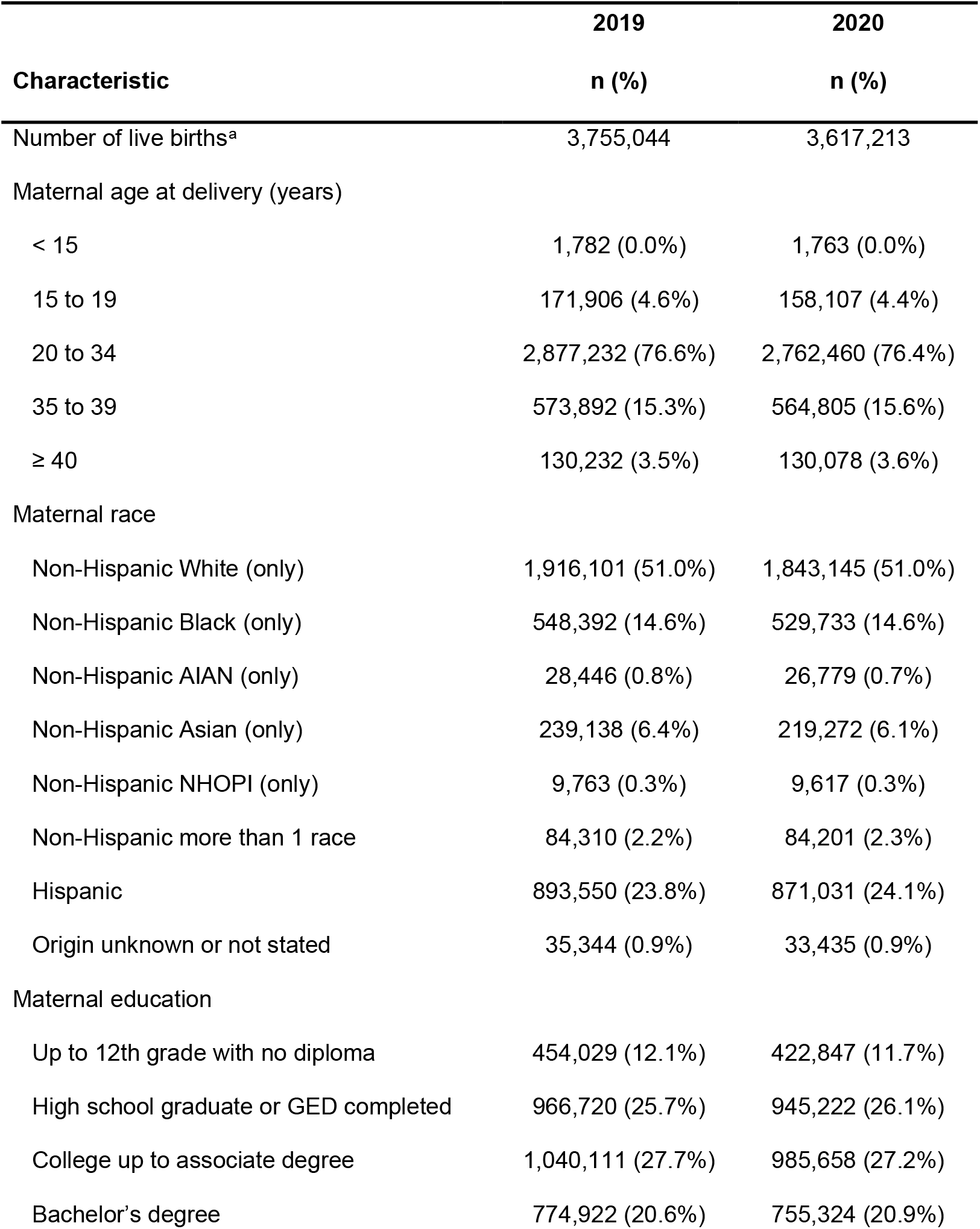
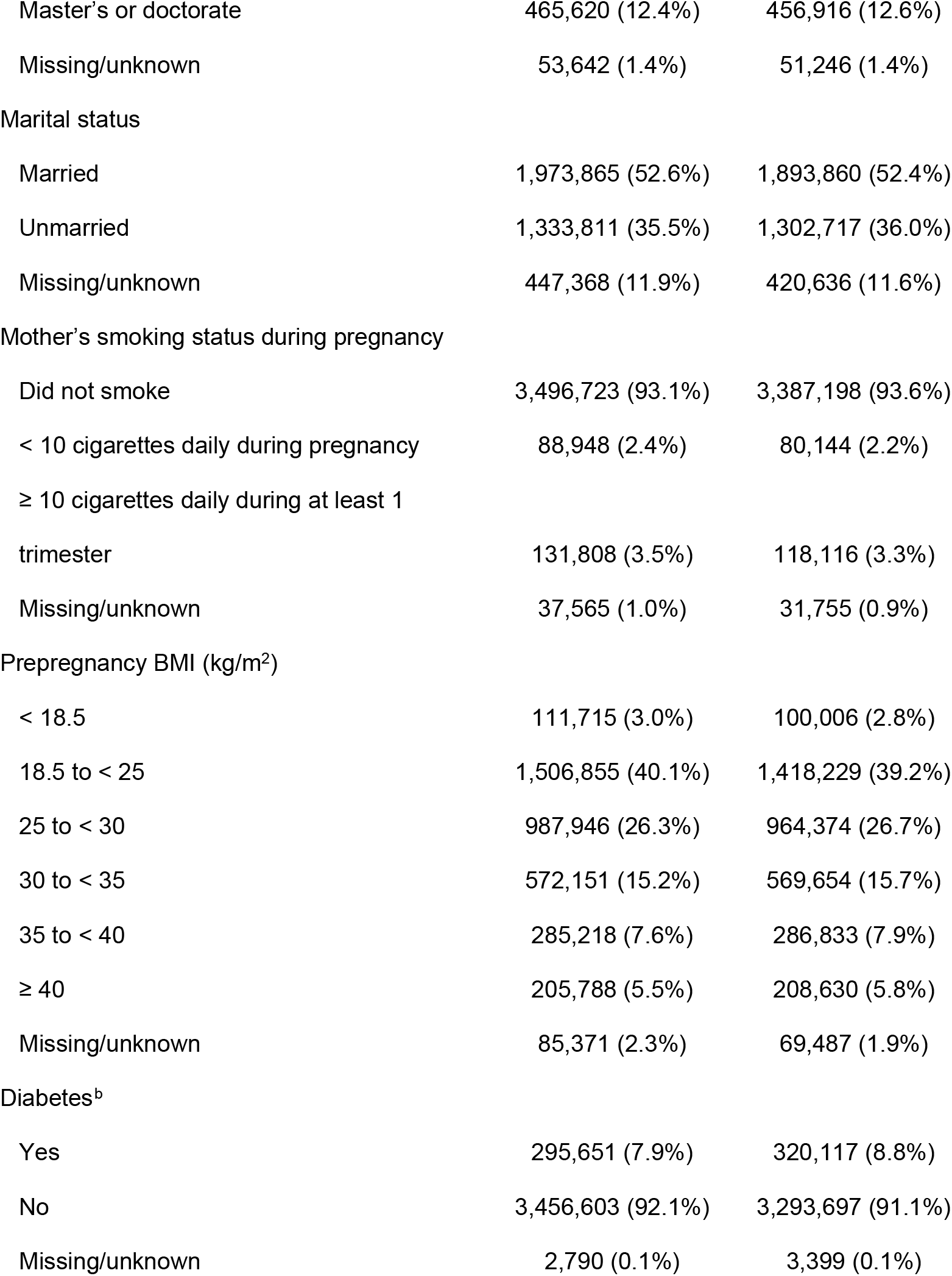
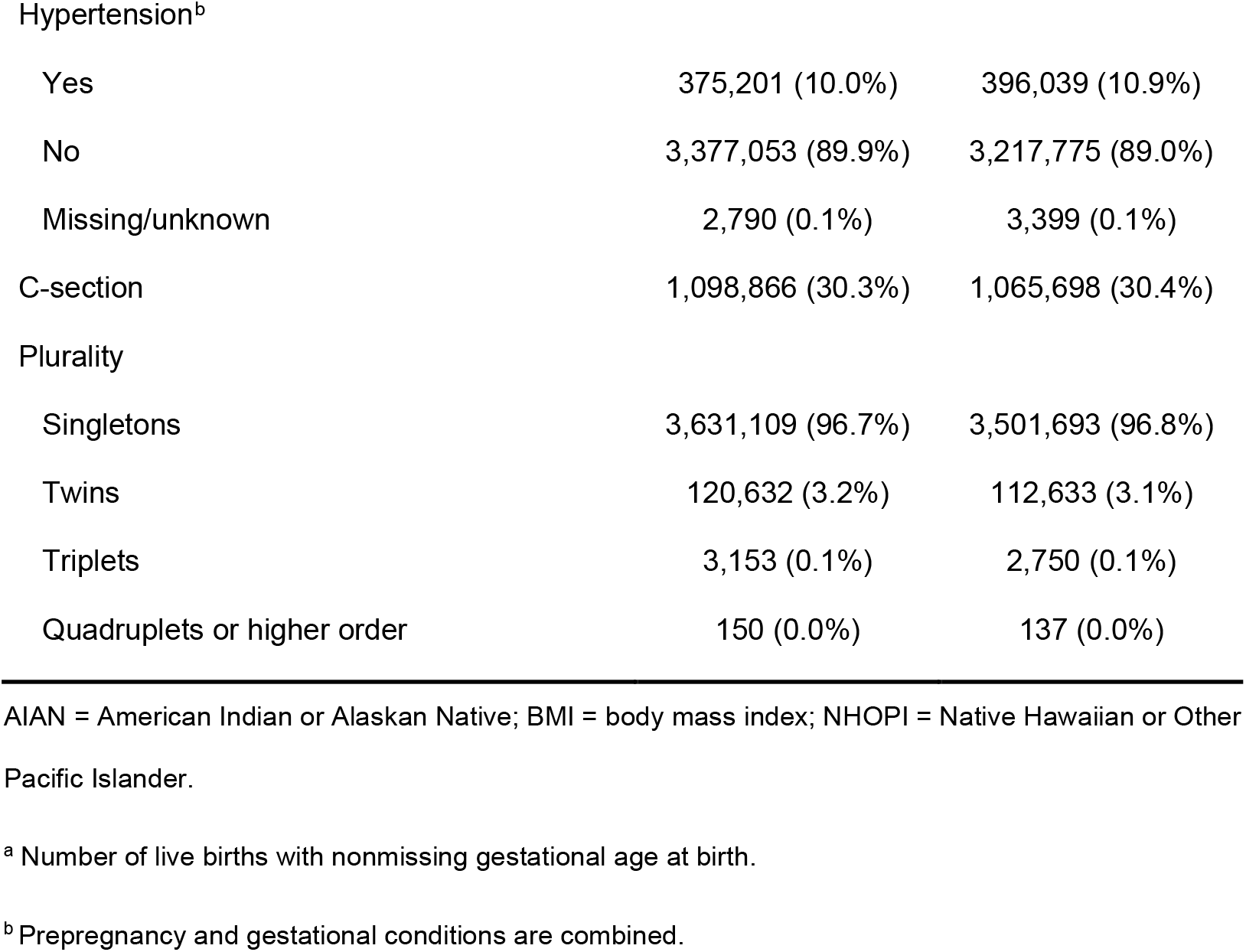
Characteristics of study population, USA 2019 and 2020.

|  | 2019 | 2020 |
| --- | --- | --- |
| Characteristic | n (%) | n (%) |
| Number of live births<sup>a</sup> | 3,755,044 | 3,617,213 |
| Maternal age at delivery (years) |  |  |
| < 15 | 1,782 (0.0%) | 1,763 (0.0%) |
| 15 to 19 | 171,906 (4.6%) | 158,107 (4.4%) |
| 20 to 34 | 2,877,232 (76.6%) | 2,762,460 (76.4%) |
| 35 to 39 | 573,892 (15.3%) | 564,805 (15.6%) |
| ≥ 40 | 130,232 (3.5%) | 130,078 (3.6%) |
| Maternal race |  |  |
| Non-Hispanic White (only) | 1,916,101 (51.0%) | 1,843,145 (51.0%) |
| Non-Hispanic Black (only) | 548,392 (14.6%) | 529,733 (14.6%) |
| Non-Hispanic AIAN (only) | 28,446 (0.8%) | 26,779 (0.7%) |
| Non-Hispanic Asian (only) | 239,138 (6.4%) | 219,272 (6.1%) |
| Non-Hispanic NHOPI (only) | 9,763 (0.3%) | 9,617 (0.3%) |
| Non-Hispanic more than 1 race | 84,310 (2.2%) | 84,201 (2.3%) |
| Hispanic | 893,550 (23.8%) | 871,031 (24.1%) |
| Origin unknown or not stated | 35,344 (0.9%) | 33,435 (0.9%) |
| Maternal education |  |  |
| Up to 12th grade with no diploma | 454,029 (12.1%) | 422,847 (11.7%) |
| High school graduate or GED completed | 966,720 (25.7%) | 945,222 (26.1%) |
| College up to associate degree | 1,040,111 (27.7%) | 985,658 (27.2%) |
| Bachelor's degree | 774,922 (20.6%) | 755,324 (20.9%) |
| Master's or doctorate | 465,620 (12.4%) | 456,916 (12.6%) |
| Missing/unknown | 53,642 (1.4%) | 51,246 (1.4%) |
| Marital status |  |  |
| Married | 1,973,865 (52.6%) | 1,893,860 (52.4%) |
| Unmarried | 1,333,811 (35.5%) | 1,302,717 (36.0%) |
| Missing/unknown | 447,368 (11.9%) | 420,636 (11.6%) |
| Mother's smoking status during pregnancy |  |  |
| Did not smoke | 3,496,723 (93.1%) | 3,387,198 (93.6%) |
| < 10 cigarettes daily during pregnancy | 88,948 (2.4%) | 80,144 (2.2%) |
| ≥ 10 cigarettes daily during at least 1 trimester | 131,808 (3.5%) | 118,116 (3.3%) |
| Missing/unknown | 37,565 (1.0%) | 31,755 (0.9%) |
| Prepregnancy BMI (kg/m<sup>2</sup>) |  |  |
| < 18.5 | 111,715 (3.0%) | 100,006 (2.8%) |
| 18.5 to < 25 | 1,506,855 (40.1%) | 1,418,229 (39.2%) |
| 25 to < 30 | 987,946 (26.3%) | 964,374 (26.7%) |
| 30 to < 35 | 572,151 (15.2%) | 569,654 (15.7%) |
| 35 to < 40 | 285,218 (7.6%) | 286,833 (7.9%) |
| ≥ 40 | 205,788 (5.5%) | 208,630 (5.8%) |
| Missing/unknown | 85,371 (2.3%) | 69,487 (1.9%) |
| Diabetes<sup>b</sup> |  |  |
| Yes | 295,651 (7.9%) | 320,117 (8.8%) |
| No | 3,456,603 (92.1%) | 3,293,697 (91.1%) |
| Missing/unknown | 2,790 (0.1%) | 3,399 (0.1%) |
| Hypertension<sup>b</sup> |  |  |
| Yes | 375,201 (10.0%) | 396,039 (10.9%) |
| No | 3,377,053 (89.9%) | 3,217,775 (89.0%) |
| Missing/unknown | 2,790 (0.1%) | 3,399 (0.1%) |
| C-section | 1,098,866 (30.3%) | 1,065,698 (30.4%) |
| Plurality |  |  |
| Singletons | 3,631,109 (96.7%) | 3,501,693 (96.8%) |
| Twins | 120,632 (3.2%) | 112,633 (3.1%) |
| Triplets | 3,153 (0.1%) | 2,750 (0.1%) |
| Quadruplets or higher order | 150 (0.0%) | 137 (0.0%) |

AIAN = American Indian or Alaskan Native; BMI = body mass index; NHOPi = Native Hawaiian or Other Pacific Islander.
<sup>a</sup> Number of live births with nonmissing gestational age at birth.
<sup>b</sup> Prepregnancy and gestational conditions are combined.

**Figure 1.**
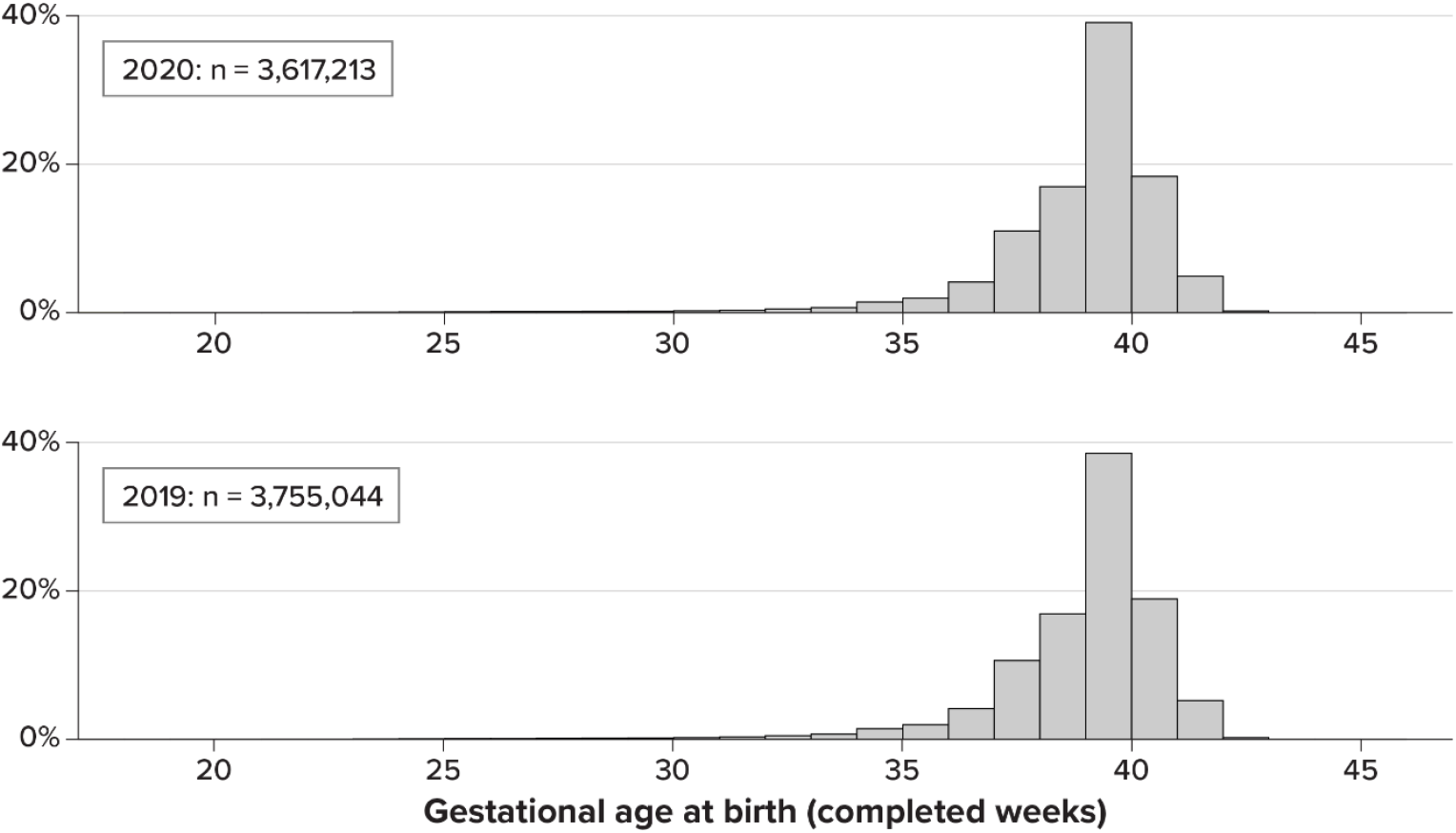
Distribution of live births by gestational age at birth, USA 2020 and 2019.

In 2020, 92.0% of live births occurred in women aged 20 to 39 years (Table 1). Overall, 51% of live births occurred in non-Hispanic White women, 24.1% in Hispanic women, and 14.6% in non-Hispanic Black women. Almost 87% of women completed high school or further studies, and 52.4% were married (11.6% had unknown marital status). Over 93% did not smoke during pregnancy; 3.3% smoked 10 or more cigarettes daily during at least 1 trimester. About 56.1% of women had BMI ≥ 25 kg/m^2^; 8.8% of women had preexisting or gestational diabetes, and 10.9% had preexisting or gestational hypertension. In 2020, 96.8% (3,501,693) of live births were singletons, 3.1% (112,633) were twins, 0.1% (2,750) were triplets, and 137 were quadruplets or higher order (Table 1; Figure 2 shows the distribution of gestational age at birth by plurality). Of all live births, 10.1% were preterm (GAB < 37 completed weeks) (median, 35 weeks; mode, 36 weeks; mean, 33.8 weeks) (Appendix B, Table B-1).

**Figure 2.**
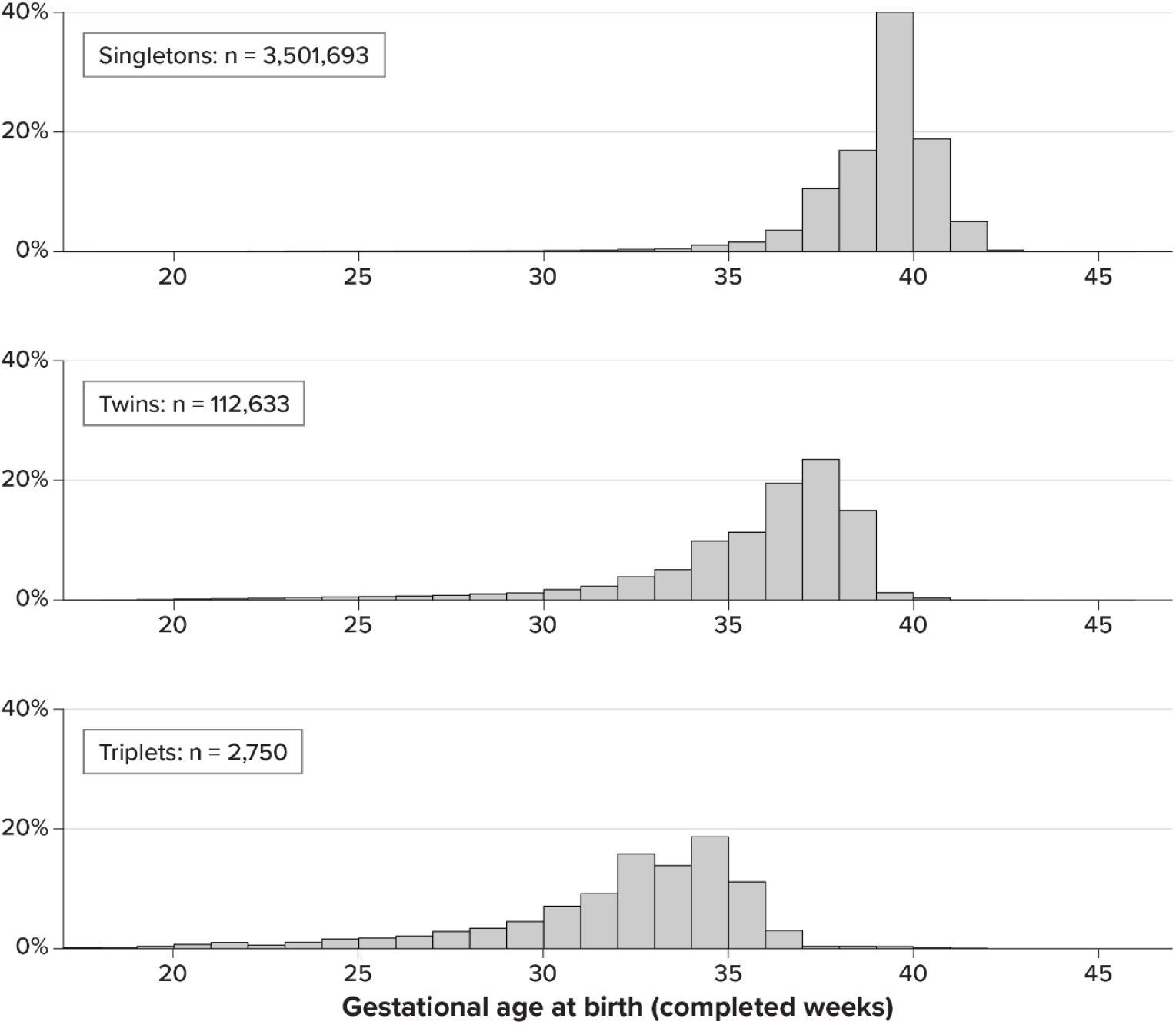
Distribution of live births by gestational age at birth, by plurality, USA 2020.

### Singletons

Among singletons (total, 3,501,693), the median and mode GAB were 39 weeks in most groups; no groups had larger median or mode GAB (Table 2; Appendix B, Table B-2); 8.4% of singletons (8.1% of live births) were preterm (median, 35 weeks; mode, 36 weeks; mean, 33.9 weeks). The following groups had lower median or mode GAB (in descending order of frequency): newborn admitted to neonatal intensive care unit (290,056 [8.3% of singletons, 8.0% of live births]; median, 37 weeks; mode, 39 weeks; mean, 35.8 weeks), low birth weight (233,500 [6.7% of singletons, 6.5% of all]; median, 35 weeks; mode, 37 weeks; mean, 34.4 weeks), very low birthweight (37,177 [1.1% of singletons, 1.0% of live births]; median and mode, 28 weeks; mean, 27.6 weeks), extremely low birthweight (17,861 [0.5% of singletons and live births]; median and mode, 25 weeks; mean, 24.9 weeks), women with eclampsia (9,263 live births [0.3% of singletons and live births], median and mode 37 weeks, mean, 36.5 weeks), newborns not discharged alive (6,730 [0.2% of singletons and live births]; median, 23 weeks; mode, 22 weeks; mean, 25.6 weeks), and women admitted to an intensive care unit as a complication of delivery or labor (5,498 [0.2% of singletons and live births]; median, 37 weeks; mode, 39 weeks; mean, 35.8 weeks).

**Table 2.**
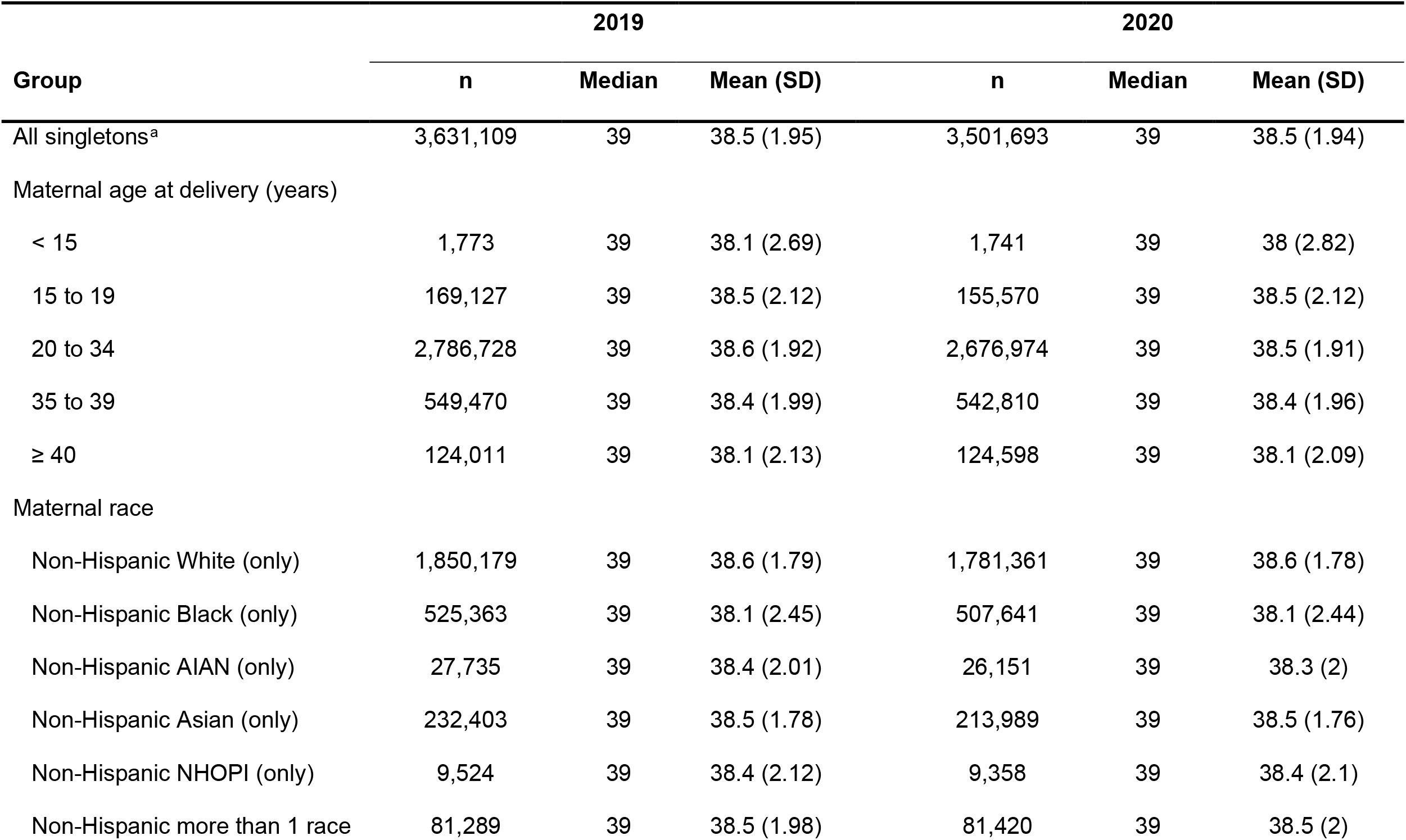
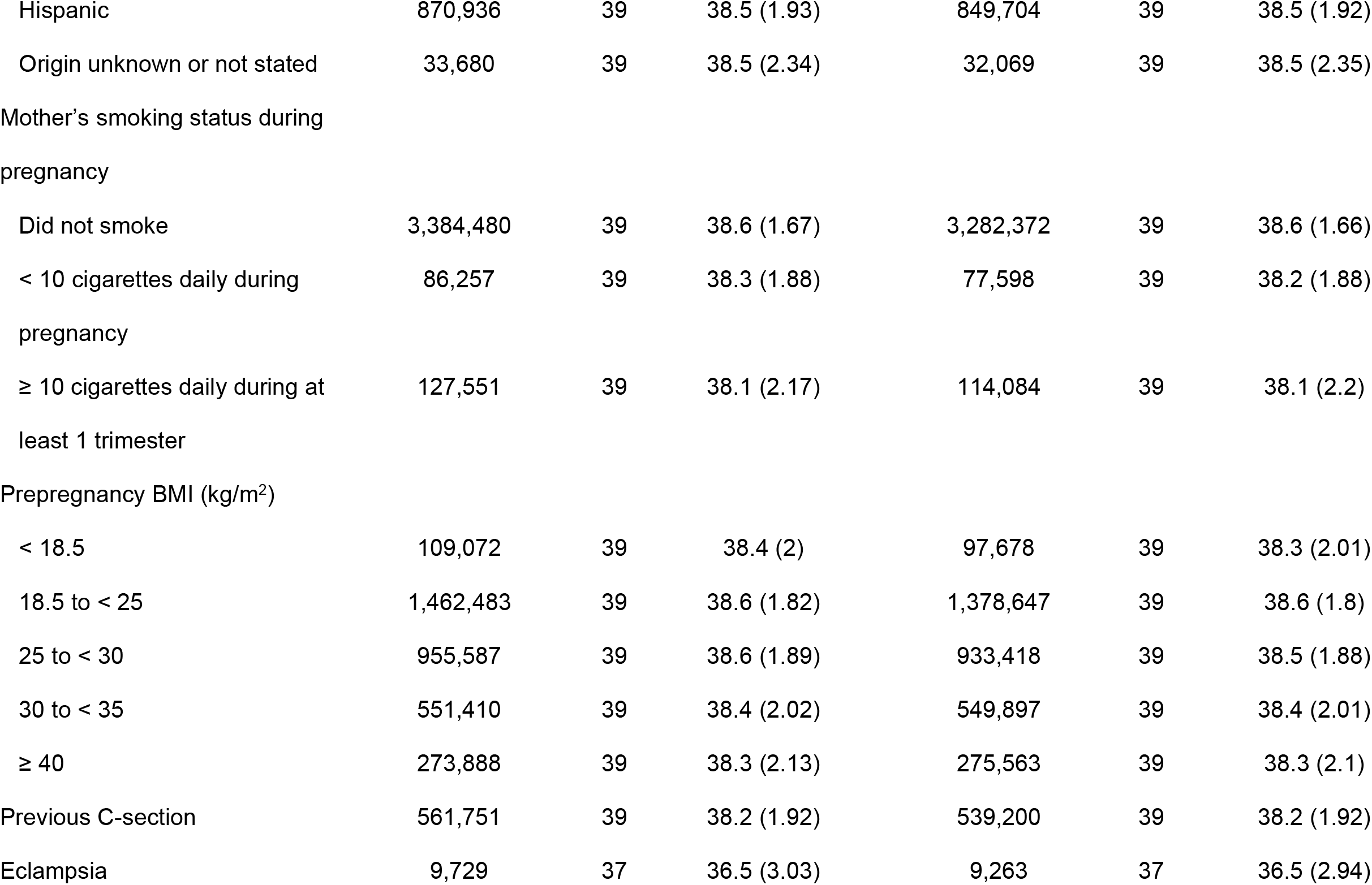
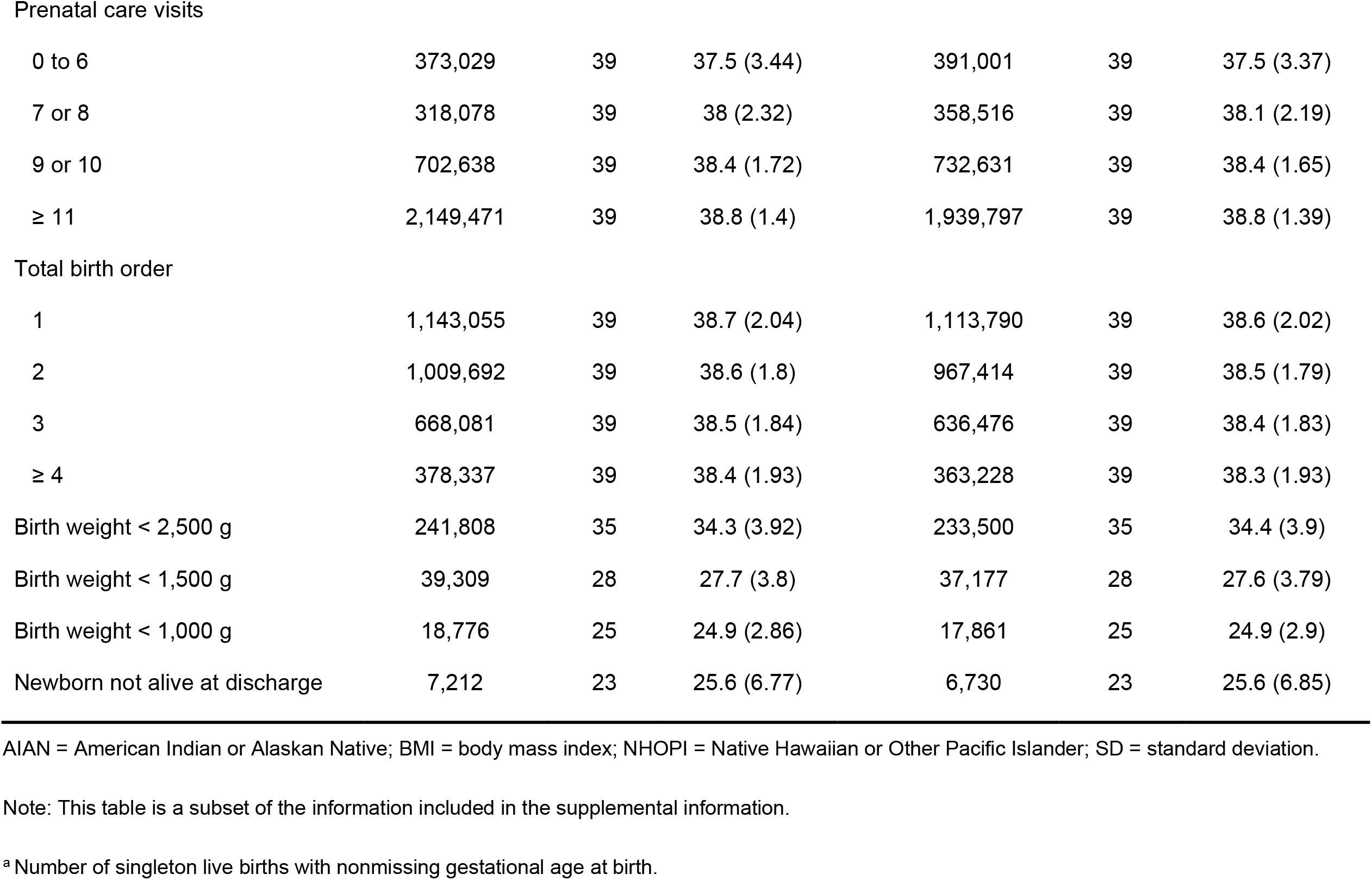
Gestational age at birth in completed weeks, singletons, USA 2019 and 2020.

| Group | 2019 |  |  | 2020 |  |  |
| --- | --- | --- | --- | --- | --- | --- |
|  | n | Median | Mean (SD) | n | Median | Mean (SD) |
| All singletons<sup>a</sup> | 3,631,109 | 39 | 38.5 (1.95) | 3,501,693 | 39 | 38.5 (1.94) |
| Maternal age at delivery (years) |  |  |  |  |  |  |
| < 15 | 1,773 | 39 | 38.1 (2.69) | 1,741 | 39 | 38 (2.82) |
| 15 to 19 | 169,127 | 39 | 38.5 (2.12) | 155,570 | 39 | 38.5 (2.12) |
| 20 to 34 | 2,786,728 | 39 | 38.6 (1.92) | 2,676,974 | 39 | 38.5 (1.91) |
| 35 to 39 | 549,470 | 39 | 38.4 (1.99) | 542,810 | 39 | 38.4 (1.96) |
| ≥ 40 | 124,011 | 39 | 38.1 (2.13) | 124,598 | 39 | 38.1 (2.09) |
| Maternal race |  |  |  |  |  |  |
| Non-Hispanic White (only) | 1,850,179 | 39 | 38.6 (1.79) | 1,781,361 | 39 | 38.6 (1.78) |
| Non-Hispanic Black (only) | 525,363 | 39 | 38.1 (2.45) | 507,641 | 39 | 38.1 (2.44) |
| Non-Hispanic AIAN (only) | 27,735 | 39 | 38.4 (2.01) | 26,151 | 39 | 38.3 (2) |
| Non-Hispanic Asian (only) | 232,403 | 39 | 38.5 (1.78) | 213,989 | 39 | 38.5 (1.76) |
| Non-Hispanic NHOPI (only) | 9,524 | 39 | 38.4 (2.12) | 9,358 | 39 | 38.4 (2.1) |
| Non-Hispanic more than 1 race | 81,289 | 39 | 38.5 (1.98) | 81,420 | 39 | 38.5 (2) |
| Hispanic | 870,936 | 39 | 38.5 (1.93) | 849,704 | 39 | 38.5 (1.92) |
| Origin unknown or not stated | 33,680 | 39 | 38.5 (2.34) | 32,069 | 39 | 38.5 (2.35) |
| Mother's smoking status during pregnancy |  |  |  |  |  |  |
| Did not smoke | 3,384,480 | 39 | 38.6 (1.67) | 3,282,372 | 39 | 38.6 (1.66) |
| < 10 cigarettes daily during pregnancy | 86,257 | 39 | 38.3 (1.88) | 77,598 | 39 | 38.2 (1.88) |
| ≥ 10 cigarettes daily during at least 1 trimester | 127,551 | 39 | 38.1 (2.17) | 114,084 | 39 | 38.1 (2.2) |
| Prepregnancy BMI (kg/m<sup>2</sup>) |  |  |  |  |  |  |
| < 18.5 | 109,072 | 39 | 38.4 (2) | 97,678 | 39 | 38.3 (2.01) |
| 18.5 to < 25 | 1,462,483 | 39 | 38.6 (1.82) | 1,378,647 | 39 | 38.6 (1.8) |
| 25 to < 30 | 955,587 | 39 | 38.6 (1.89) | 933,418 | 39 | 38.5 (1.88) |
| 30 to < 35 | 551,410 | 39 | 38.4 (2.02) | 549,897 | 39 | 38.4 (2.01) |
| ≥ 40 | 273,888 | 39 | 38.3 (2.13) | 275,563 | 39 | 38.3 (2.1) |
| Previous C-section | 561,751 | 39 | 38.2 (1.92) | 539,200 | 39 | 38.2 (1.92) |
| Eclampsia | 9,729 | 37 | 36.5 (3.03) | 9,263 | 37 | 36.5 (2.94) |
| Prenatal care visits |  |  |  |  |  |  |
| 0 to 6 | 373,029 | 39 | 37.5 (3.44) | 391,001 | 39 | 37.5 (3.37) |
| 7 or 8 | 318,078 | 39 | 38 (2.32) | 358,516 | 39 | 38.1 (2.19) |
| 9 or 10 | 702,638 | 39 | 38.4 (1.72) | 732,631 | 39 | 38.4 (1.65) |
| ≥ 11 | 2,149,471 | 39 | 38.8 (1.4) | 1,939,797 | 39 | 38.8 (1.39) |
| Total birth order |  |  |  |  |  |  |
| 1 | 1,143,055 | 39 | 38.7 (2.04) | 1,113,790 | 39 | 38.6 (2.02) |
| 2 | 1,009,692 | 39 | 38.6 (1.8) | 967,414 | 39 | 38.5 (1.79) |
| 3 | 668,081 | 39 | 38.5 (1.84) | 636,476 | 39 | 38.4 (1.83) |
| ≥ 4 | 378,337 | 39 | 38.4 (1.93) | 363,228 | 39 | 38.3 (1.93) |
| Birth weight < 2,500 g | 241,808 | 35 | 34.3 (3.92) | 233,500 | 35 | 34.4 (3.9) |
| Birth weight < 1,500 g | 39,309 | 28 | 27.7 (3.8) | 37,177 | 28 | 27.6 (3.79) |
| Birth weight < 1,000 g | 18,776 | 25 | 24.9 (2.86) | 17,861 | 25 | 24.9 (2.9) |
| Newborn not alive at discharge | 7,212 | 23 | 25.6 (6.77) | 6,730 | 23 | 25.6 (6.85) |

AIAN = American Indian or Alaskan Native; BMI = body mass index; NHOPi = Native Hawaiian or Other Pacific Islander; SD = standard deviation.
Note: This table is a subset of the information included in the supplemental information.
<sup>a</sup> Number of singleton live births with nonmissing gestational age at birth.

### Twins

Among twins in 2020 (total, 112,633), median GAB was 36 weeks and mode was 37 weeks in most groups (Appendix B, Table B-3); 59.9% of twins (1.9% of live births) were preterm (median, 35 weeks; mode, 36 weeks; mean, 33.5 weeks). As with singletons, no groups had a larger median or mode GAB. The characteristics that identified groups with lower median or mode GAB among singletons also did with twins. Additional groups that had lower median or mode GAB were (in descending frequency) pregnancies with 0 to 6 prenatal visits (15,530 live births [13.8% of twins, 0.4% of live births]; median, 34 weeks; mode, 36 weeks; mean, 32.5 weeks), with 7 or 8 prenatal visits (13,013 live births [11.6% of twins, 0.4% of live births]; median, 35 weeks; mode, 36 weeks; mean, 34.3 weeks), pregnancies in which the mother smoked 10 or more cigarettes per day during pregnancy (3,965 [3.5% of twins, 0.1% of live births]; median and mode, 36 weeks; mean, 34.6 weeks), with maternal age 15 to 19 years (2,499 live births [2.2% of twins, 0.1% of live births]; median, 35 weeks; mode, 37 weeks; mean, 34.1 weeks), in which the mother smoked 1 to 9 cigarettes per day during pregnancy (2,497 [2.2% of twins, 0.1% of live births]; median and mode, 36 weeks; mean 35.0 weeks), and with maternal age < 15 years (19 live births [0.02% of twins, 0.001% of all live births]; median, 35 weeks; mode, 36 weeks; mean, 32.2 weeks. As among singletons, newborns not discharged alive had the smallest median and mode GAB.

### Other results

Generally, pregnancies with characteristics that can be considered healthy had a narrower GAB distribution; for example, 77.8% of singletons of women with BMI between 18.5 and less than 25 kg/m^2^ had GAB within 1 week around the mode (38 through 40 weeks), while only 35.9% of pregnancies in which newborns were admitted into the neonatal intensive care unit had GABs within 1 week around the mode (also 38 through 40 weeks). Newborns with birthweight < 1,500 grams had practically the same GAB distribution, regardless of whether they were singletons or twins, and these distributions were broader than that for singletons with birthweight ≥ 1,500 grams (Appendix A, Figure A-1; Appendix B, Table B-3). Among newborns not discharged alive, GAB had a mode at 21 weeks (13.2% of 8,162 newborns) and a small increase at 37 weeks (3.7%) (Appendix A, Figure A-2, and Appendix B, Table B-1).

The means displayed more variation than the medians and modes; SDs often increased as means decreased. For example, the mean (SD) GAB was 38.6 (1.7) weeks for singletons of women who did not smoke during pregnancy in 2020; 38.2 (1.9) weeks for women who smoked < 10 cigarettes daily during pregnancy, and 38.1 (2.2) weeks for women who smoked 10 or more cigarettes daily in at least 1 trimester; however, the median and mode were 39 weeks for these 3 groups (Table 2; Appendix B, Table B-2).

Mean squared errors were smaller when calculated using the mean than when using the median or mode; in contrast, mean absolute values of the errors were smaller when calculated using the median than when using the mean or the mode (when the median and mode were the same, mean absolute values of the errors were smaller when calculated using them than when calculated using the mean; Appendix B, Table B-4).

## DISCUSSION

### Main results

Birth certificates from the US in 2019 and 2020 indicated that newborns overall and in most groups defined by maternal and newborn characteristics had a median and mode GAB of 39 weeks; this was driven by the GAB in singletons (96.8% of pregnancies with nonmissing GAB). Among singletons, live births—including live births in women of any age, who smoked or did not smoke during pregnancy, with any BMI—had a median and mode GAB of 39 weeks; median or mode GAB lower than 39 weeks was observed in pregnancies with complications in the mother or the offspring. Multifetal pregnancies had lower GAB: for twins, overall and in several groups, the median was 36 weeks and the mode was 37 weeks; GAB was shorter for triplets. Among twins, additional groups with lower median or mode GAB were identified in groups based on number of prenatal care visits, maternal age, and smoking during pregnancy. We observed larger variability of GAB across groups in twins than in singletons.

### How can these results be useful?

Data sources with valuable medication exposure information may lack some pregnancy-specific information; an example is US claims data sources, which are often used for perinatal pharmacoepidemiologic research. When individual-level clinical or obstetric estimates of duration of pregnancy are lacking, researchers often use a fixed number of weeks to estimate pregnancy duration and date of pregnancy start, to then ascertain the timing of drug or other exposures relative to the start of pregnancy. Our results can be used in several ways in this process. First, our results can be used to refine pregnancy-identifying or pregnancy-dating algorithms, allowing researchers to identify smaller groups for more individualized GAB estimation based on characteristics of each pregnancy or newborn that may be available in their data source. For example, a singleton pregnancy with known eclampsia could be assigned 37 weeks (mode and median, 2020), instead of 39 weeks (overall mode and median, 2020), thus reducing misclassification of exposure. Second, our results can be used to probabilistically impute 1 GAB value for each pregnancy as a random draw from the appropriate GAB distribution. For example, a singleton pregnancy in a 30-year old woman would have a 16.8% probability of being imputed a GAB of 38 weeks; 39.9%, 39 completed weeks (the mode); 19.5%, 40 weeks, etc. (2020 data). The multiple imputation version of this process would further reflect the uncertainty around the duration of pregnancies in the face of missing information.

Researchers could draw various GAB values per pregnancy (producing, for example, 10 completed data sets), conduct all the downstream analyses for each completed data set and finally combine the results. In addition, we provide current GAB distributions and information on which statistic can reduce errors in GAB estimates.

US birth files have 1 observation per liveborn infant: multifetal pregnancies are represented multiple times. Despite this, our results are applicable to studies whose unit of observation is pregnancies because we provide results for singletons and, separately, for twins, and because GAB distributions (percentages by gestational week, mean, median, and mode) are the same for newborns and for the corresponding pregnancies within each of those groups (assuming all twins are born alive).

Research groups have used the median^1,2,4^ or the mode^1^ GAB to estimate pregnancy duration; one may wonder whether the median, the mode, or even the mean GAB is most appropriate for estimations. We found that the median and the mode are the same for many groups. The mean squared distance, which penalizes large differences, always favored using the mean over the median or the mode. On the other hand, the mean absolute value of the distance always favored the median (and the mode, when they were the same). Researchers wanting to minimize the number of days (i.e., linear distance) between the imputed and the true value, based on our results, could use the median GAB; researchers wanting to minimize the squared distance could use the mean GAB.

Our results highlight that most singleton groups had a median and mode GAB of 39 weeks, including groups determined by BMI and smoking, data that may not be available in the claims data sources often used for perinatal pharmacoepidemiology research. While some groups with lower GAB were small (e.g., live births from women with eclampsia were 0.3% of all births), they may be the target of interventions and research, as these groups often were pregnancies with maternal or newborn complications.

### Generalizability

Our results are generalizable to subpopulations within the US and to populations elsewhere with similar healthcare practices; for example, in England, the mode GAB has been reported as 39 weeks (29.1% of live births with known duration of pregnancy), with 68% of deliveries taking place from 38 to 40 completed weeks in April 2020 through March 2021^13^ (compared with 38.8% live births at 39 weeks and 73.9% born at 38 to 40 weeks in the US in 2020). In populations where healthcare practices differ considerably from the US, the distribution of GAB might vary. For example, 32% of pregnancies were estimated to result in cesarean section in North America in 2018 (also observed in the data that we used in 2019 and 2020; Appendix B, Table B-1), but cesarean sections comprised 43% of pregnancies in Latin America and the Caribbean and 16% in Southeast Asia.^14^ Temporal changes in GAB in the US have been documented, with the most common duration of singleton livebirth pregnancies with spontaneous delivery shifting from 40 weeks in 1992 to 39 weeks in 2002;^15^ our analyses show that 39 weeks was still the mode (and also median) GAB among all live births and live births born via cesarean section in the US in 2020.

### Missing data

The lack of information on pregnancy start date or GAB can be seen as a missing data problem. Using information from maternal and infant characteristics to estimate GAB (such as using group-specific GAB distributions), as we propose, makes the missing-at-random assumption more plausible.^16^ Our proposal to use information obtained after delivery/birth to estimate GAB (e.g., using information on whether the mother or the newborn received intensive care) is appropriate because what happens downstream from an unobserved event contains information on the unobserved event (as wet cars in the street can be an indication of unobserved earlier rain) and is consistent with established multiple imputation approaches.^17^

### Strengths and limitations

For these analyses, we used a very large population-based data source that contains information on GAB and maternal and newborn characteristics. This allowed us to explore combinations of variables and still have a large number of observations within groups. Furthermore, US birth certificates have been found to be a valid source of information on duration of pregnancy or GAB^18-20^ and have been used as gold standard in validating claims-based algorithms estimating GAB.^21-23^ However, they have been reported as not reliable for other data elements such as maternal weight,^24^ smoking during pregnancy,^25^ and other characteristics.^26^ Birth weight, mode of delivery, and presence of some maternal chronic conditions have also been found to be reliable,^18,26^ and linkage to birth certificates has been advocated for research on drug safety in pregnancy in healthcare databases.^27^ Another strength of our study is that our results might be used to mitigate misclassification of GAB among shorter pregnancies, a known limitation of some previous research.^21,28-30^

Limitations of this study include the aforementioned birth certificate shortcomings and the fact that US birth files include only live births. Despite the large size of the data source, the number of triplets and higher order multifetal pregnancies was small, and we did not explore them separately. For similar practical reasons, we explored only 2-variable combinations. In the original data source, GAB is presented in completed weeks; finer granularity is not provided. Some characteristics that identify groups with lower GAB, such as birth weight, may not be available in some data sources.

## CONCLUSIONS

Most singleton live births, including live births in women of any age, who smoked or did not smoke during pregnancy, and with any BMI had a median and mode GAB of 39 weeks. Some live birth groups had distinct GAB distributions; these groups can be identified from characteristics recorded in many existing data sources used for observational epidemiologic research. GAB distributions provided here can be useful in estimating GAB when clinical estimates are not available in those data sources.

## Data Availability

We used US birth data files of the Centers for Disease Control and Prevention (CDC) for years 2019 (the most recent year before the COVID-19 pandemic) and 2020 (the most recent available data). These files are publicly available for download.
References with links:
CDC. US Centers for Disease Control and Prevention. Vital statistics online data portal: downloadable data files. 13 May 2022. https://www.cdc.gov/nchs/data_access/vitalstatsonline.htm. Accessed 15 May 2022.
CDC. US Centers for Disease Control and Prevention. Guide to completing the facility worksheets for the certificate of live birth and report of fetal death (2003 revision, update September 2019). September 2019. https://www.cdc.gov/nchs/data/dvs/GuidetoCompleteFacilityWks.pdf. Accessed 23 December 2021.

https://www.cdc.gov/nchs/data_access/vitalstatsonline.htm

## ACKNOWLEDGEMENTS

Editorial services were provided by Adele Monroe, ELS, and graphic art services were provided by Bethan Pickering, both employees of RTI Health Solutions. We would like to thank Abenah Harding, also an employee of RTI Health Solutions, for her help preparing this manuscript. Development of this manuscript was supported financially by RTI Health Solutions.

### Appendix A. Additional information

This is supplemental information for the paper

#### Checklist for reporting in perinatal pharmacoepidemiology

**Table A-1.**
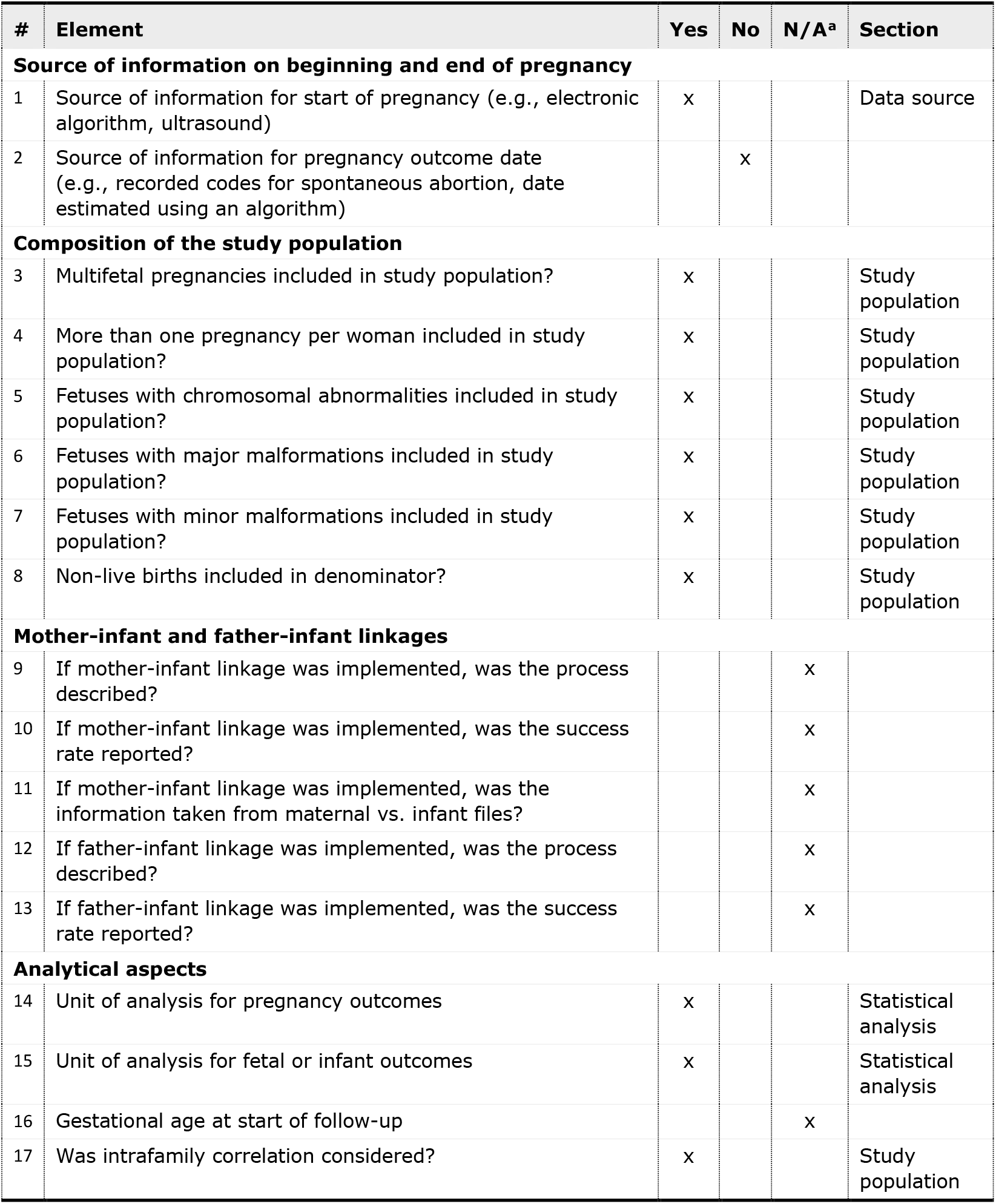
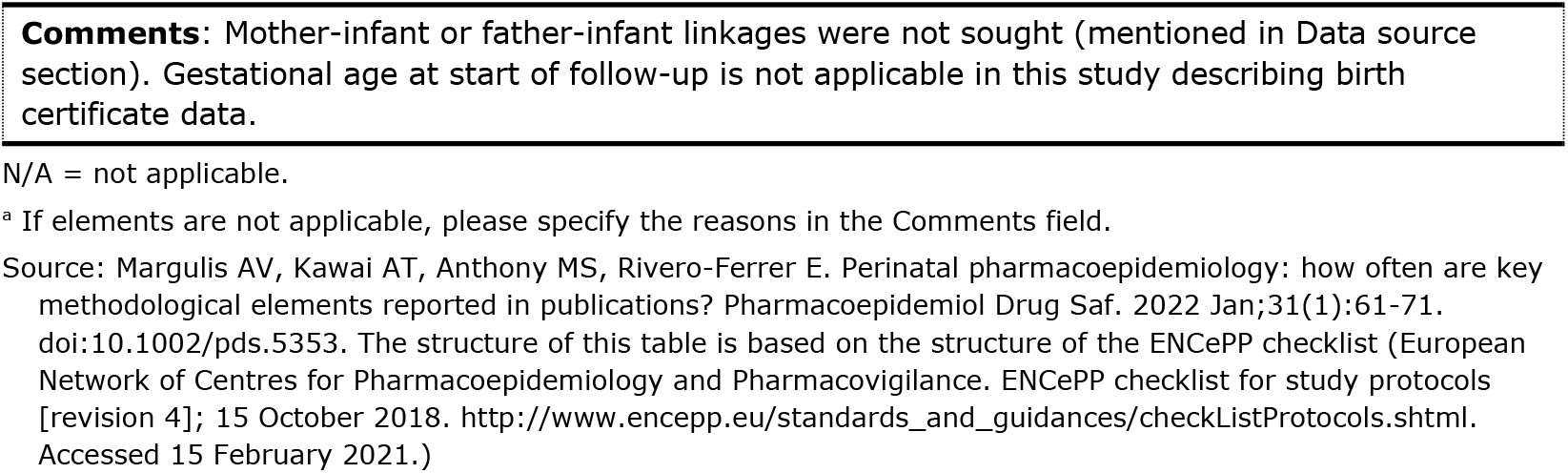
Checklist for reporting in perinatal pharmacoepidemiology.

#### Study variables

**Table A-2.**
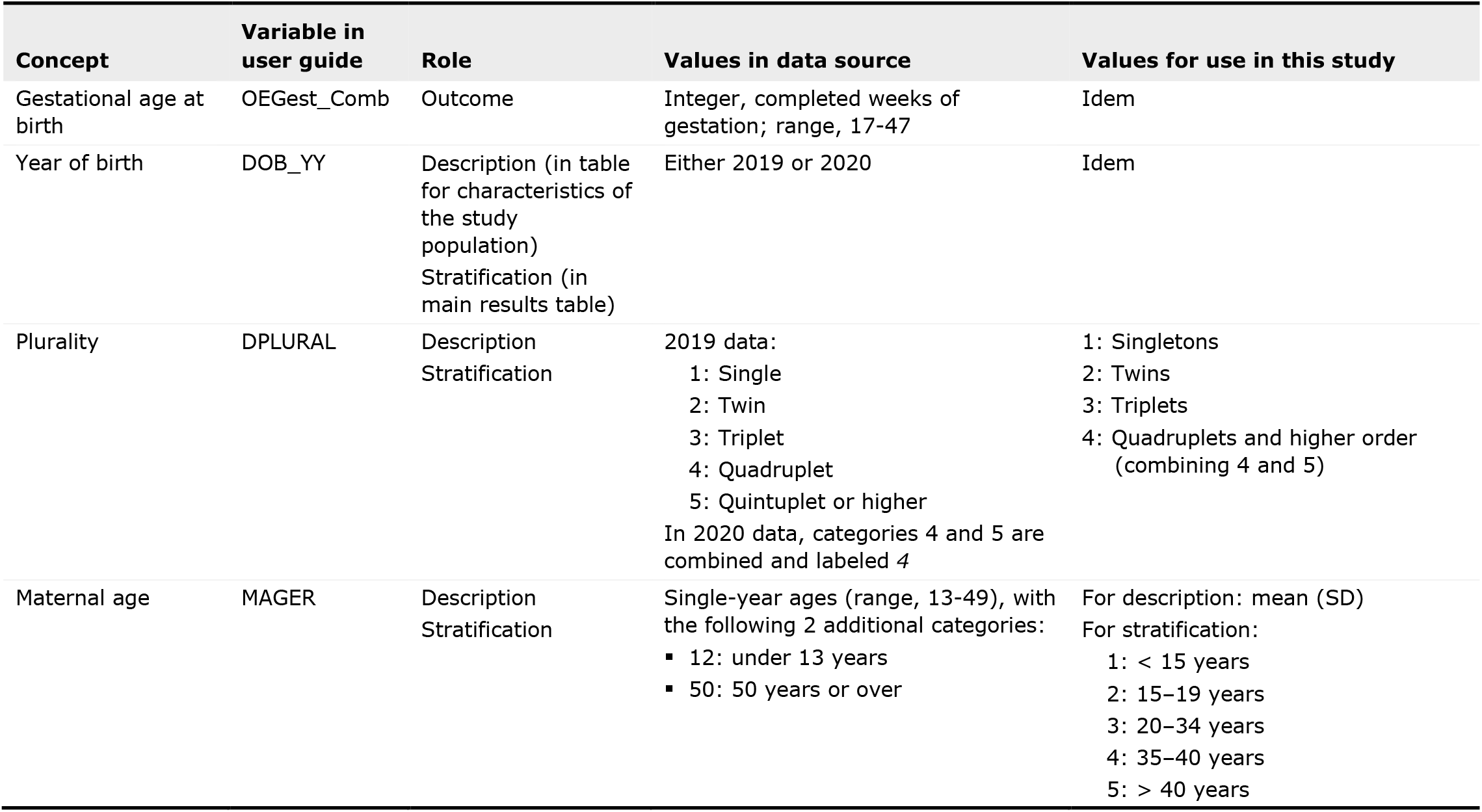
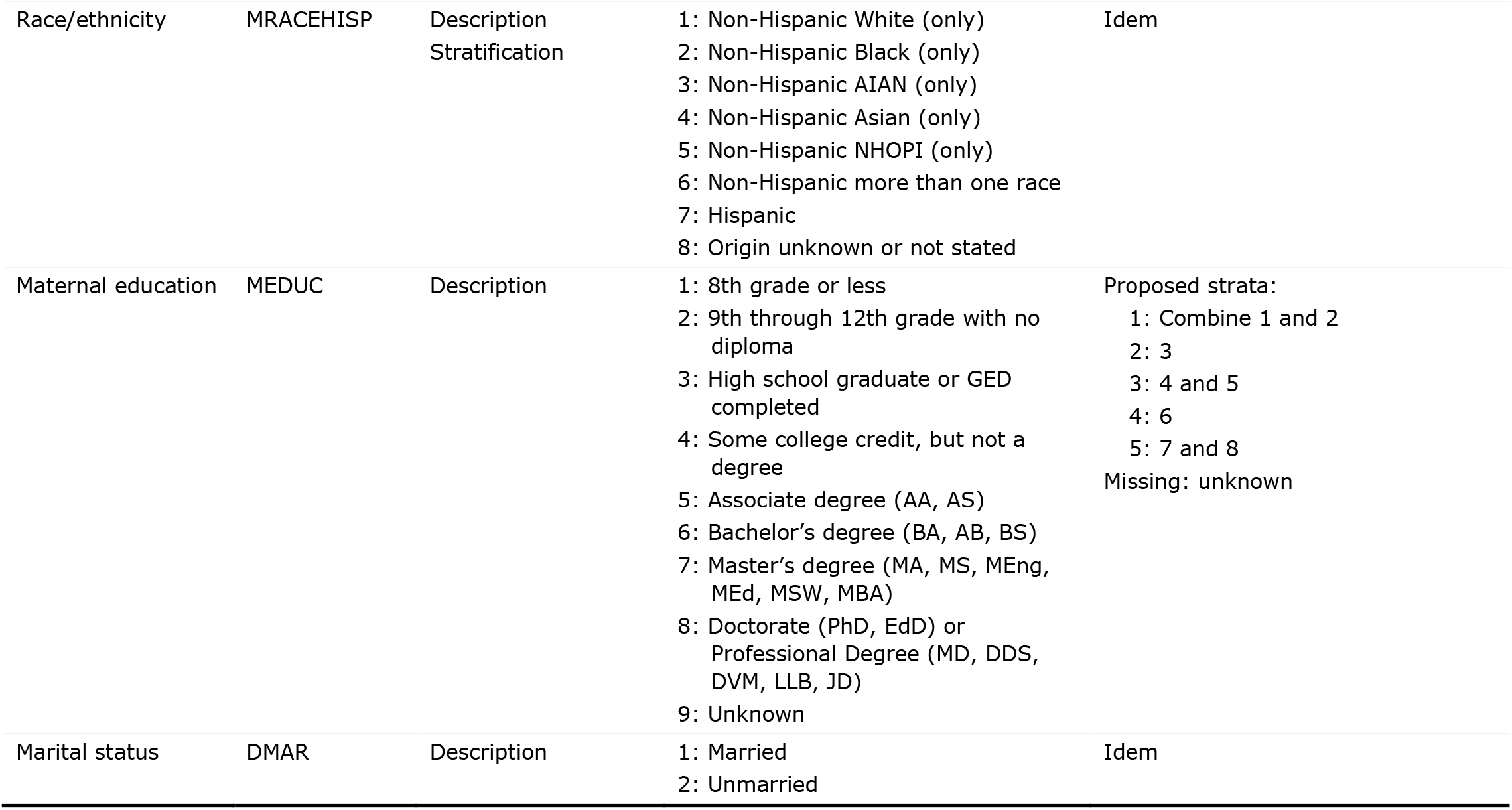
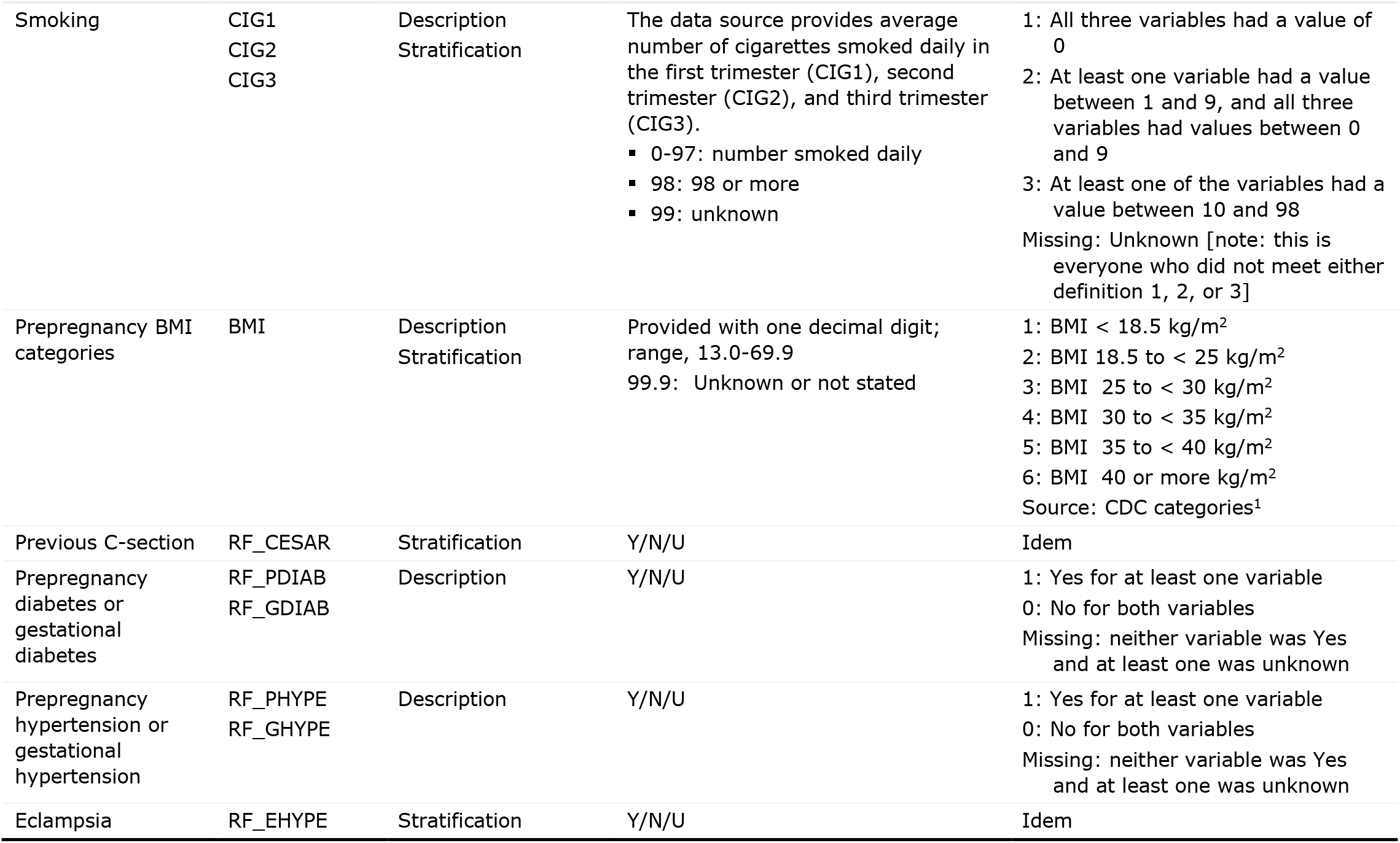
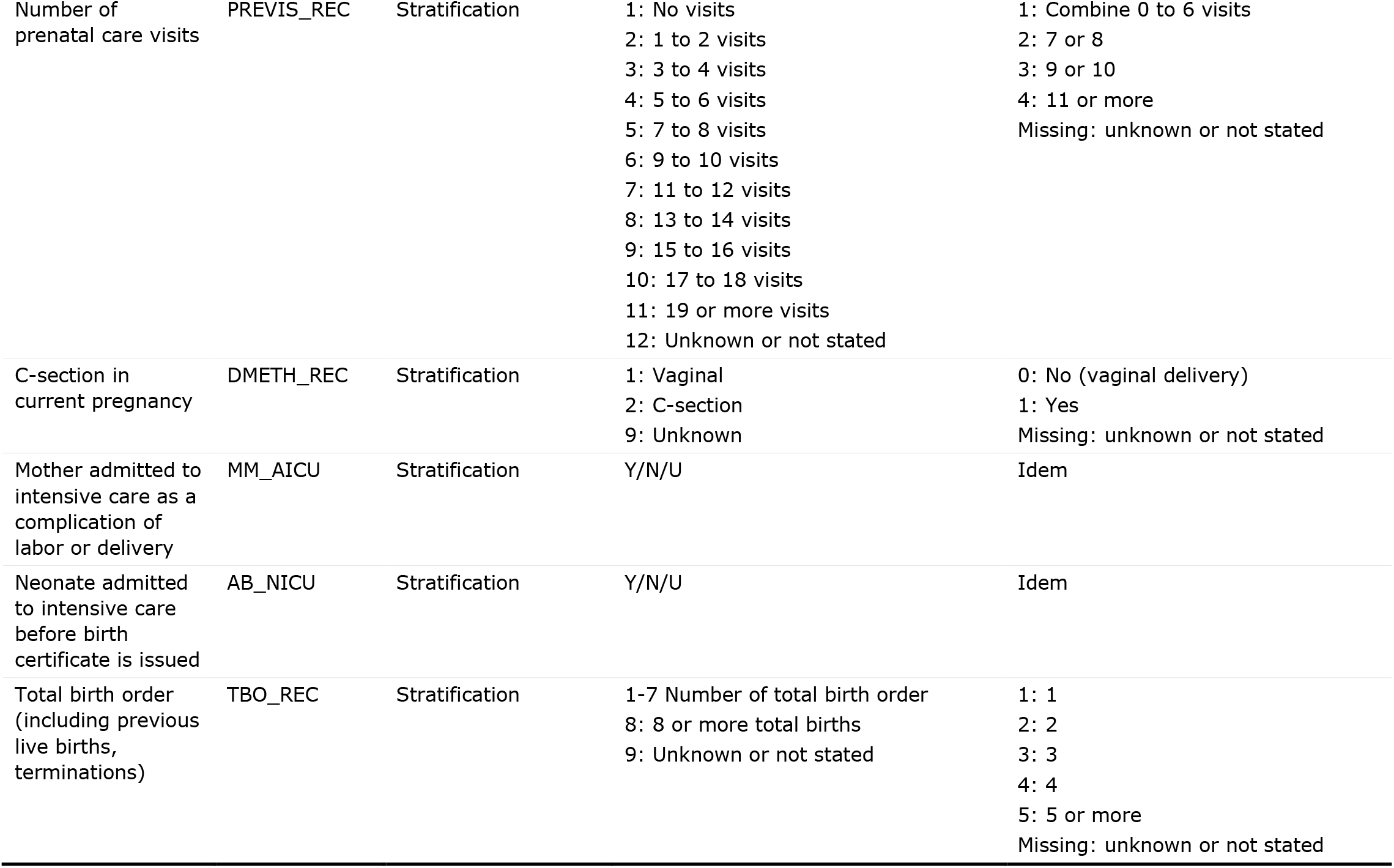
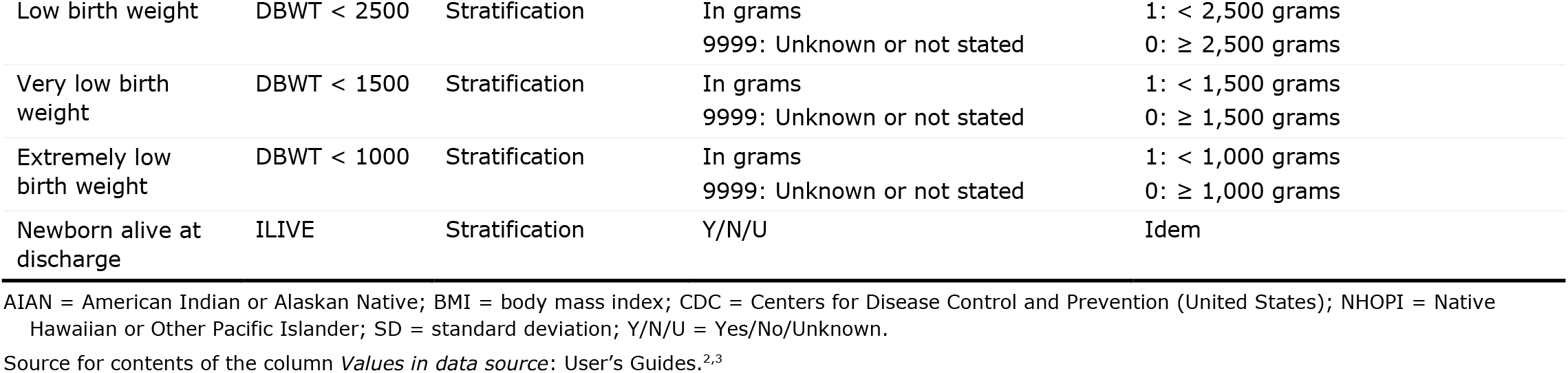
Study variables.

### Methods to explore whether the median, mode, or mean would result in a smaller error in estimating gestational age at birth

Analyses in this study were aimed at informing the estimation of gestational age at birth or duration of pregnancy in other data sources. One way of doing this is, using an example, to assign all singletons born small for gestational age whose gestational age is unknown the median gestational age observed in 2019 in the present study. To support recommendations on which summary statistic should be used (i.e., median, mode, or mean), we calculated two values:

1. The *mean squared error* for subgroups. The mean squared error using the median was calculated as follows: 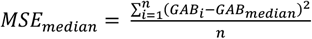 for every observation *i* in the group of size *n* In words, following the same example: the mean squared error using the median was the gestational age at birth for live birth *i* (singleton born small for gestational age) minus the median gestational age at birth among singletons born small for gestational age, squared, averaged across all singletons born small for gestational age. This mean squared error was calculated for the median, mode, and mean. A smaller mean squared error reflects a better estimation.
2. The *mean absolute value of the error* for selected subgroups: 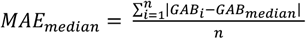 for every observation *i* in the group of size *n* In words, following the same example: the mean absolute error using the median was the absolute value of gestational age at birth for live birth *i* (singleton born small for gestational age) minus the median gestational age at birth among singletons born small for gestational age, averaged across all singletons born small for gestational age. This statistic was calculated for the median, mode, and mean. A smaller value reflects a better estimation.

#### Additional figures

**Figure A-1.**
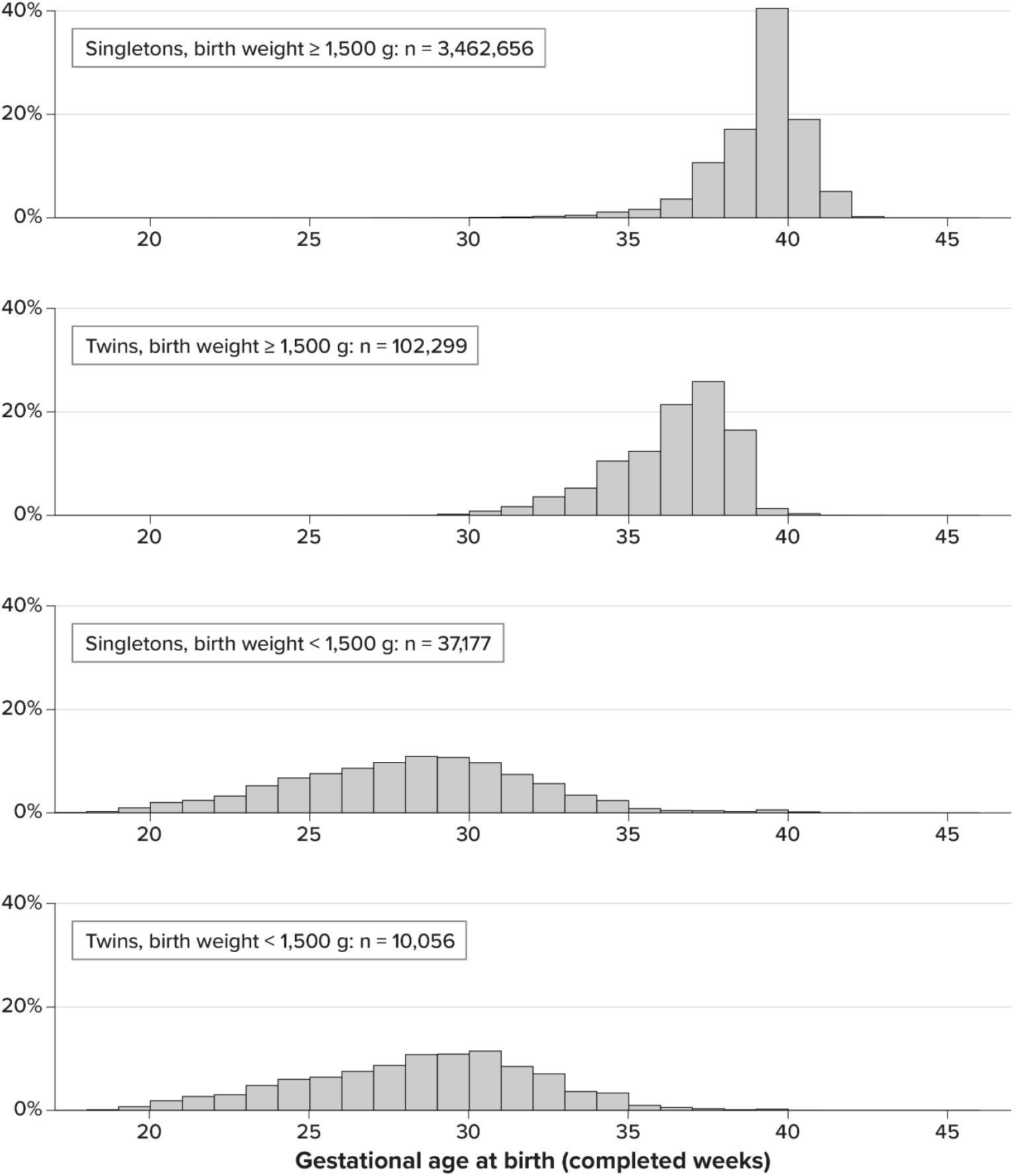
Distribution of live births by gestational age at birth by plurality and birth weight, USA 2020.

**Figure A-2.**
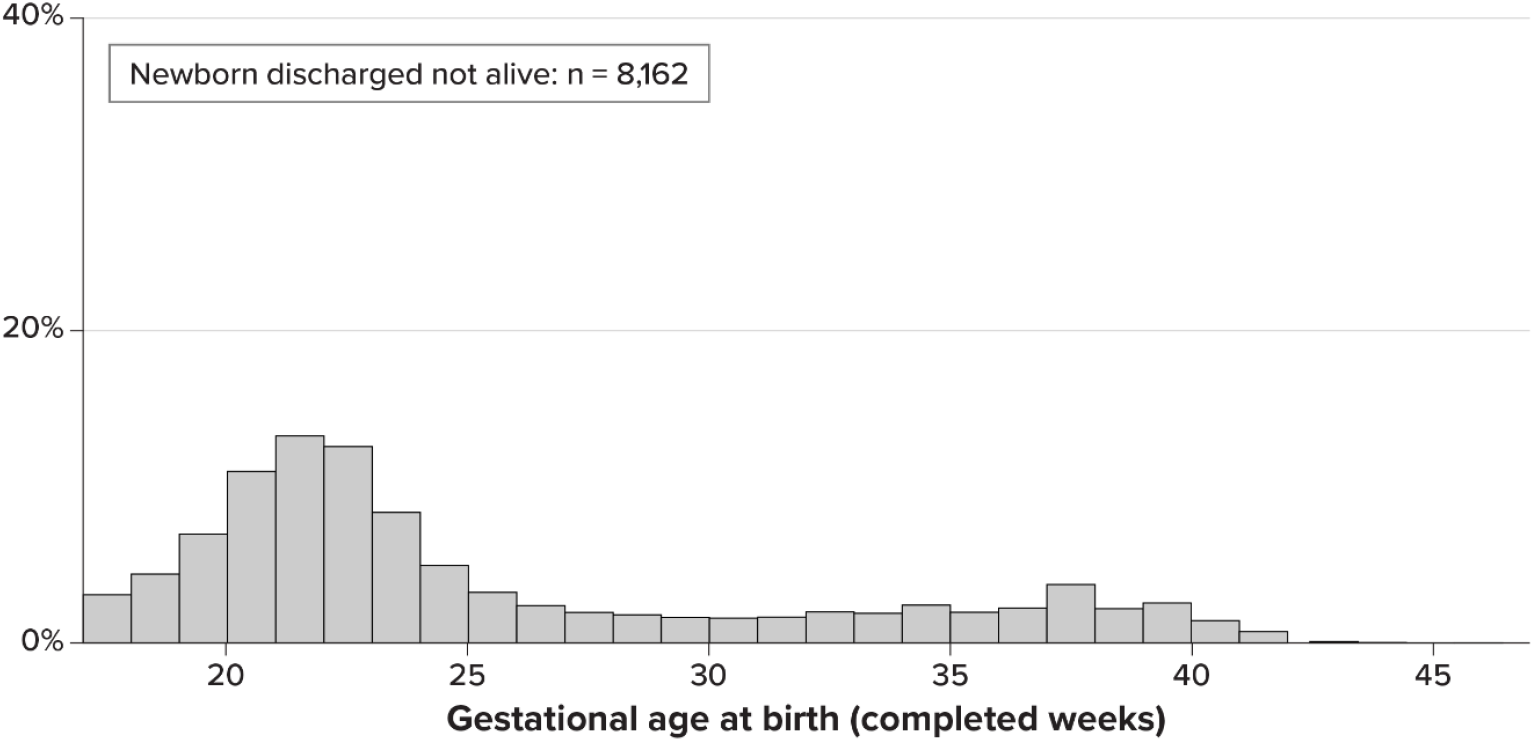
Distribution live births by gestational age at birth in newborns not discharged alive, USA 2020.

## Notes

**Funding Statement:** This work was supported by RTI Health Solutions.

**CONFLICT(S) OF INTEREST** The authors have no conflict of interest for this publication

### Competing Interest Statement

The authors have declared no competing interest.

### Funding Statement

This work was supported by RTI Health Solutions.

### Author Declarations

We used US birth data files of the Centers for Disease Control and Prevention (CDC) for years 2019 (the most recent year before the COVID-19 pandemic) and 2020 (the most recent available data). These files are publicly available for download. References with links: CDC. US Centers for Disease Control and Prevention. Vital statistics online data portal: downloadable data files. 13 May 2022. https://www.cdc.gov/nchs/data_access/vitalstatsonline.htm. Accessed 15 May 2022. CDC. US Centers for Disease Control and Prevention. Guide to completing the facility worksheets for the certificate of live birth and report of fetal death (2003 revision, update September 2019). September 2019. https://www.cdc.gov/nchs/data/dvs/GuidetoCompleteFacilityWks.pdf. Accessed 23 December 2021.

